# Pre-diabetes as a Critical Stage for Risk of Dementia and Stroke: Evidence from the UK Biobank and Mendelian Randomization

**DOI:** 10.1101/2025.09.30.25336879

**Authors:** Si Han, Elnaz Naderi, Kan Wang, Yuan Ma, Geert Jan Biessels, Fariba Ahmadizar

## Abstract

**Background and Aims:** Type 2 diabetes (T2D) is a recognized risk factor for dementia and stroke, but whether risks begin during pre-diabetes remains unclear. We aimed to determine the risks of these neurological conditions with pre-diabetes and the glycemic thresholds at which risks emerge.

**Methods:** We studied 432,285 adults (mean age, 57 years; 55% women) from the UK Biobank who were free from prior dementia or stroke at baseline. Glycemic status (normoglycemia, pre-diabetes, T2D) was defined by HbA1c. Incident dementia, stroke, and MRI-derived brain markers, including brain volumes and white matter hyperintensity volumes, were assessed for these conditions. HbA1c was modeled continuously to assess non-linear effects. Outcomes were estimated using competing risks and linear regression models, adjusted for demographic, lifestyle, and clinical factors. Two-sample Mendelian randomization (MR) was used to test causality.

**Results:** During a median 13.7 years of follow-up, pre-diabetes was associated with higher risks of Alzheimer’s disease (HR:1.30; 95% CI: 1.16-1.45), vascular dementia (HR:1.50; 95% CI: 1.28-1.7), and all-cause stroke (HR:1.24; 95% CI: 1.16-1.33), independent of other vascular risk factors. On brain MRI (N=3,723), total and grey matter volumes and hippocampal volumes were smaller, and white matter hyperintensity volume was larger in pre-diabetes. Spline models indicated that adverse outcomes emerged at HbA1c levels around 40-45 mmol/mol, below the diabetes threshold. Risks were further elevated in T2D. MR analyses supported causal effects of glycemia on Alzheimer’s disease, stroke, and hippocampal atrophy.

**Conclusion:** Pre-diabetes is not a benign state: even modest HbA1c elevations are linked to dementia, stroke, and brain structural decline. These risks emerge below current diagnostic glycemic thresholds, underscoring pre-diabetes as a critical stage for early intervention to preserve brain health.

**Lay Summary:** This study shows that even before diabetes develops, higher HbA1c levels are linked to increased risks of dementia, stroke, and brain damage.

- People with pre-diabetes had a higher chance of developing Alzheimer’s disease, other forms of dementia, such as vascular dementia, and stroke. Brain scans also showed earlier signs of shrinkage and damage.
- These risks appeared even before blood sugar reached the level used to diagnose diabetes, highlighting the importance of early monitoring and healthy lifestyle changes to reduce the risk of dementia and stroke.

## Introduction

Type 2 diabetes mellitus (T2D) is a major global health challenge, affecting more than 530 million people worldwide, a number projected to reach 783 million by 2045 (1). Yet the impact of dysglycemia extends beyond diagnosed diabetes. Pre-diabetes, defined by modest elevations in hemoglobin A1c (HbA1c) or fasting glucose, is highly prevalent, affecting an estimated 1 in 3 adults globally (2, 3). While often considered a “pre-disease” state, emerging evidence suggests that pre-diabetes may carry its own risk of adverse outcomes.

Neurological disorders represent a particularly urgent concern. Dementia is projected to affect over 150 million people by 2050 (4), and stroke remains one of the leading causes of death and disability worldwide (5). Both conditions share vascular and metabolic risk factors, and stroke is a well-established contributor to subsequent cognitive decline (6, 7). Identifying modifiable upstream factors that increase risk of both stroke and dementia is therefore a major public health priority.

The role of T2D in increasing dementia and stroke risk is well established (8–15).

However, whether risks begin earlier, during pre-diabetes or even within the “high-normal” glycemic range, remains uncertain. Some studies have linked pre-diabetes to a higher risk of dementia and stroke (16, 17), while others reported null associations (18, 19).

To address these gaps, we examined associations of glycemic status and HbA1c with dementia, stroke, and MRI-derived brain markers in the UK Biobank. Recognizing the limitations of observational analyses, we complemented these findings with Mendelian randomization (MR) to strengthen causal inference (20). By integrating large-scale cohort data with genetic approaches, our study aimed to determine whether pre-diabetes is already associated with adverse neurological outcomes and to identify the glycemic thresholds at which risk becomes apparent.

## Methods

### 2.1 Study design and data sources

We conducted a prospective cohort analysis using data from the UK Biobank (UKB), a population-based resource of over 500,000 participants aged 40-69 years recruited between 2006 and 2010. Participants completed questionnaires, underwent interviews, and physical assessments, and provided biological samples at 22 centers across England, Scotland, and Wales. Outcomes were ascertained through linkage to hospital admissions, death registries, primary care records, and algorithm-derived dementia classifications. Ethical approval was obtained from the Northwest Multi-centre Research Ethics Committee (16/NW/0274), and all participants provided written informed consent (21). We excluded individuals with dementia or stroke at baseline, those with missing HbA1c, and those first diagnosed with pre-diabetes or T2D after the baseline visit. To minimize reverse causation, dementia and stroke events were included only if diagnosed at least two years after the recorded onset of glycemic abnormality. The final analytic sample and the imaging subsample are shown in *Figure 1*.

**Figure 1.**
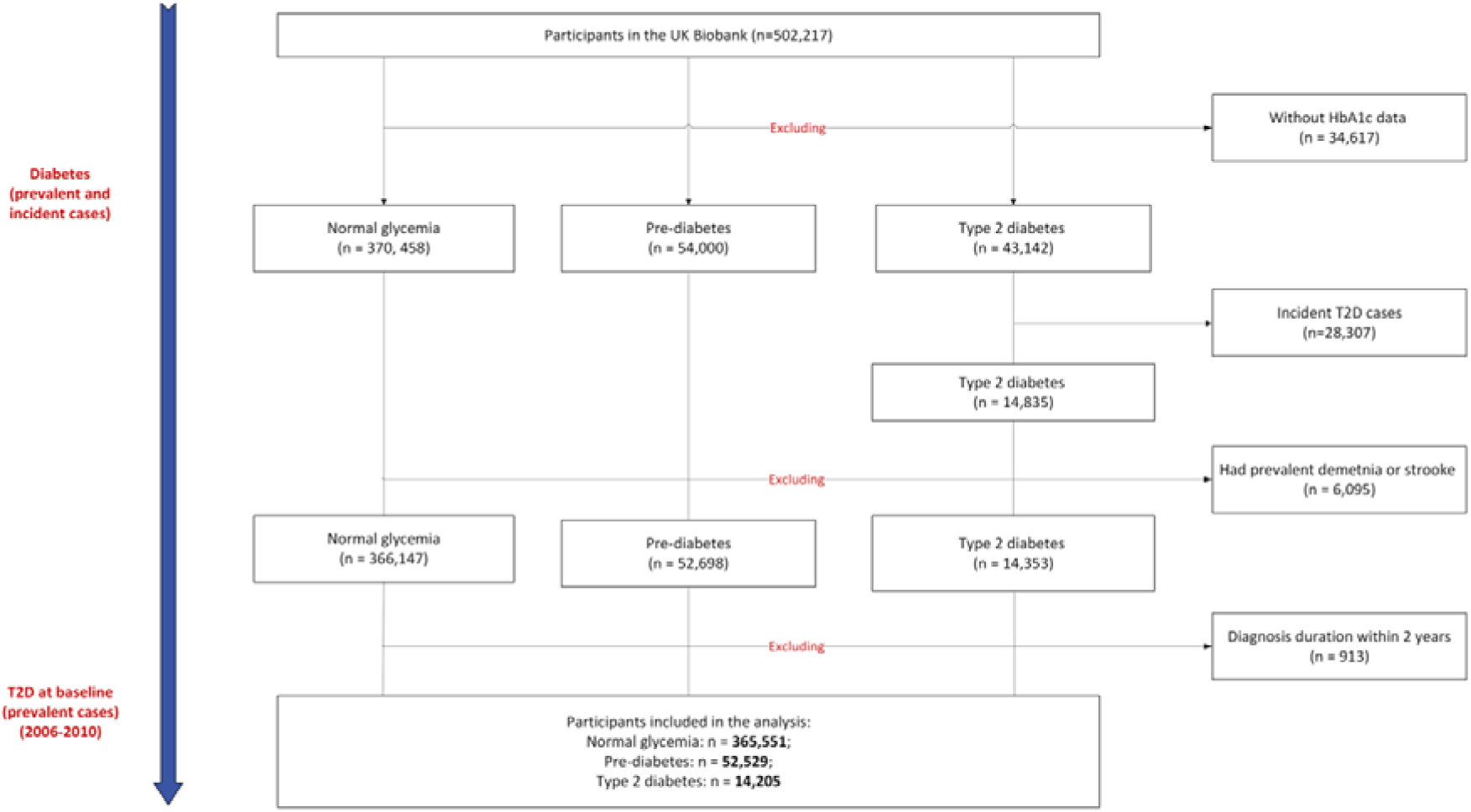
Flow diagram of study participants. Flow chart showing the selection of participants from the UK Biobank cohort, including exclusions, baseline classification of glycemic status (normal glucose, pre-diabetes, type 2 diabetes), and follow-up outcomes. Participants with missing or unreliable HbA1c data, incident type 2 diabetes after baseline, prior prevalent diagnoses, or diagnosis within two years of baseline were excluded.

### 2.2. Exposures

The primary exposure was glycemic status at baseline, classified as: normoglycemia: HbA1c <5.7% (<39 mmol/mol); pre-diabetes: HbA1c 5.7-6.4% (39-47 mmol/mol) and without using glucose lowering medications, and T2D: documented diagnosis prior to baseline (self-report or health records) or undiagnosed diabetes meeting ADA criteria (HbA1c ≥6.5% [≥48 mmol/mol] or random glucose ≥11.1 mmol/L). Undiagnosed cases labeled “abnormal glucose” in UKB were classified as T2D, given the adult age range. Full definitions and coding algorithms are provided in the *Supplementary Figure S1*. To capture dose-response across the glycemic spectrum, HbA1c was also modeled continuously using restricted cubic splines.

### 2.2 Outcomes

Primary outcomes were incident dementia (all-cause, Alzheimer’s disease [AD], vascular dementia [VaD]) and stroke (all-cause, ischemic, intracerebral hemorrhage), defined using hospital, primary care, and death records. Secondary outcomes were MRI-derived brain structural measures (total brain volume, grey and white matter volumes, hippocampal volume, and white matter hyperintensity [WMH] volume) available in an imaging subsample.

Outcome code lists and UKB data-field mappings are provided in the *Supplementary Material S1*.

### 2.3 Covariates

Covariates were selected based on known associations with cognitive, vascular, and metabolic health. Demographic factors included age, sex, ethnicity, education, and socioeconomic deprivation. Lifestyle factors included smoking status, alcohol consumption, dietary habits, and physical activity. Clinical covariates included body mass index (BMI), blood pressure, family history of diabetes and dementia, and medication use (lipid-lowering, blood pressure-lowering, and glucose-lowering drugs). Genetic variables included *APOE* ε4 status and a polygenic risk score (PRS) for T2D. Variable definitions are provided in the *Supplementary Material S2*.

### 2.4 Statistical methods

Baseline characteristics were summarized across glycemic categories. For time-to-event outcomes, we used Fine-Gray competing-risks regression to estimate SHRs and 95% confidence intervals (CIs), treating death as a competing event. Time at risk was from baseline to the first qualifying outcome, death, or censoring at last follow-up. Participants with outcome diagnoses on/before baseline were excluded. For individuals with pre-diabetes or T2D, we restricted to those whose diagnosis occurred on or before baseline. To minimize reverse causation, outcome events within two years of the recorded onset of glycemic abnormality were excluded.

Models were progressively adjusted for demographic and lifestyle factors, followed by clinical/treatment factors, and finally genetic factors (model components in *Figure 2*). Cumulative incidence functions were plotted by glycemic category; group differences were assessed using Gray’s test.

**Figure 2.**
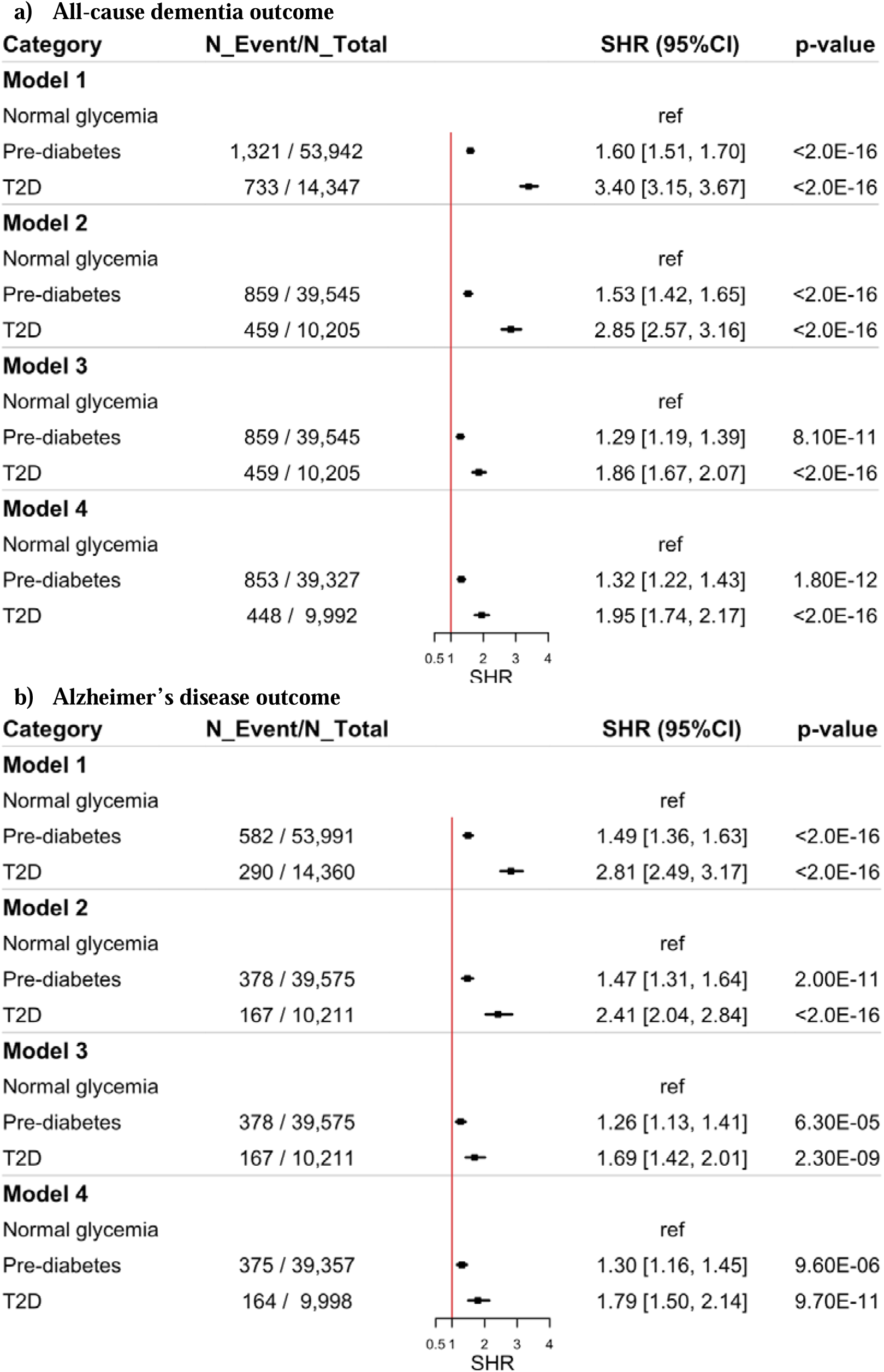
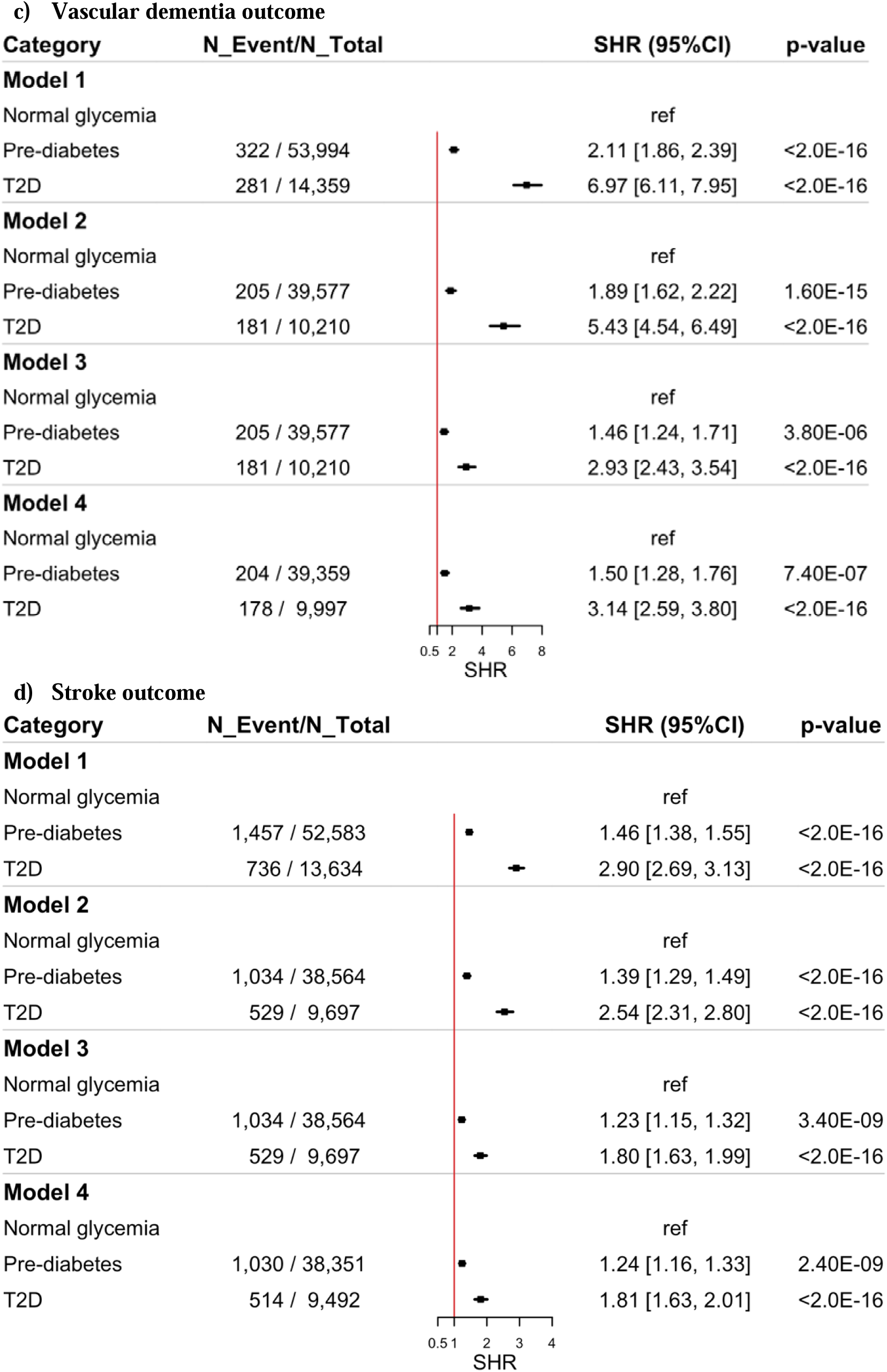
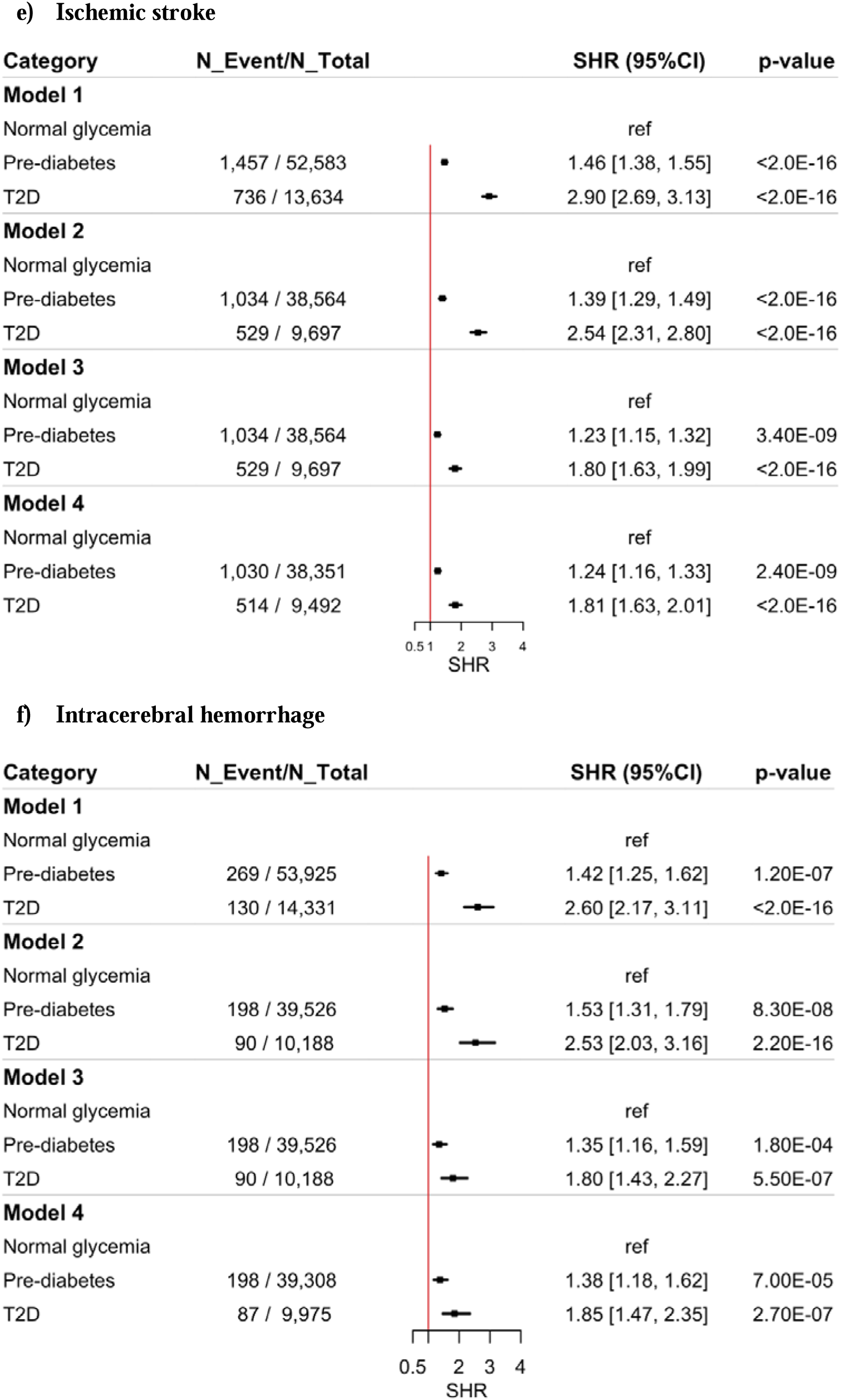
(a-f). Subdistribution hazard ratios for dementia and stroke outcomes associated with pre-diabetes and type 2 diabetes across four adjustment models. Abbreviations: T2D, type 2 diabetes; SHR, subdistribution hazard ratio; CI, confidence interval; FDR, false discovery rate. Model adjustments: Model 1 is crude. Model 2 is adjusted for age, gender, ethnicity, education, deprivation, diet, smoking, alcohol, and BMI. Model 3 further includes family history of diabetes, family history of dementia, and medication use of blood pressure-lowering medications, lipid-lowering medications, oral glucose-lowering medications, and insulin. Model 4 adds adjustments for T2D polygenic risk score and APOE4 carrier status.

MRI outcomes were analyzed using linear regression of standardized measures (z-scores), of which WMH volume was first log-transformed (log1p) to reduce skewness, then standardized. Results are reported as standardized β coefficients with 95% confidence intervals. HbA1c was modeled continuously with restricted cubic splines to evaluate non-linear associations for both clinical and MRI outcomes.

To control for multiple comparisons across MRI outcomes, we applied Benjamini-Hochberg false discovery rate (FDR) correction. Multicollinearity was assessed using variance inflation factors (VIFs); diagnostics indicated no covariate exceeded accepted thresholds. All analyses were performed using R version 4.5.1 (2025-06-13) (R Foundation for Statistical Computing, Vienna, Austria).

### 2.5 Mendelian Randomization analyses

Two-sample MR was used to assess the causal effects of genetically predicted T2D and HbA1c on dementia, stroke, and MRI phenotypes. Genome-wide significant single-nucleotide polymorphisms (SNPs) were selected as instruments, pruned for linkage disequilibrium, and screened for pleiotropy. Steiger filtering was applied to reduce reverse causation, and only SNPs with F-statistics >10 was retained. Primary analyses used inverse-variance weighting (IVW), with sensitivity analyses conducted using MR-Egger, weighted median, and MR-PRESSO. To examine independence from adiposity, multivariable MR (MVMR) adjusting for genetically predicted BMI was performed. Bonferroni correction was applied to account for multiple testing. Further technical details are provided in the *Supplementary Materials (Table S1, Material S3-S4)*.

## Results

### 3.1 Study Population and Baseline Characteristics

At baseline, participants with pre-diabetes (N = 52,529) were older than those with normal glycemia (median age 61 vs 57 years). The median HbA1c was 40.4 mmol/mol in those with pre-diabetes, 34.3 mmol/mol in those with normal glycemia, and 51.7 mmol/mol in participants with T2D. Compared with normoglycemia, the pre-diabetes group had a higher prevalence of obesity (33% vs 19%) and hypertension (31% vs 17%), and greater lipid-lowering therapy use (32% vs 16%). Lifestyle patterns also differed: fewer reported daily alcohol intake (37% vs 46%) and more were current smokers (15% vs 9.5%). Overall, pre-diabetes showed intermediate clinical/metabolic characteristics between normal glycemia and T2D. All between-group differences were statistically significant (p<0.001). Full characteristics are in *Table 1*.

**Table 1.** Baseline sociodemographic, clinical, and lifestyle characteristics stratified by glycemic status: normal glycemia, pre-diabetes, and type 2 diabetes.

| Variable | Overall<br>N = 432,285 | Normal glycemia<br>N = 365,551 | Pre-diabetes<br>N = 52,529 | T2D<br>N = 14,205 | p-value |
| --- | --- | --- | --- | --- | --- |
| **Age (median (Q1, Q3))** | 57 (50, 63) | 57 (49, 62) | 61 (55, 65) | 61 (55, 65) | <0.001 |
| **Sex (n, (%))** |  |  |  |  | <0.001 |
| Female | 238,522 (55%) | 203,784 (56%) | 29,376 (56%) | 5,362 (38%) |  |
| Male | 193,763 (45%) | 161,767 (44%) | 23,153 (44%) | 8,843 (62%) |  |
| **Ethnicity (n, (%))** |  |  |  |  | <0.001 |
| Asian | 8,736 (2.0%) | 5,671 (1.6%) | 2,057 (3.9%) | 1,008 (7.1%) |  |
| Black | 5,430 (1.3%) | 3,368 (0.9%) | 1,612 (3.1%) | 450 (3.2%) |  |
| Others | 5,988 (1.4%) | 4,531 (1.2%) | 1,104 (2.1%) | 353 (2.5%) |  |
| White | 410,170 (95%) | 350,511 (96%) | 47,423 (90%) | 12,236 (86%) |  |
| **Education (n, (%))** |  |  |  |  | <0.001 |
| College/University degree | 143,459 (33%) | 125,860 (34%) | 14,434 (27%) | 3,165 (22%) |  |
| Other | 288,826 (67%) | 239,691 (66%) | 38,095 (73%) | 11,040 (78%) |  |
| **Deprivation (n, (%))** |  |  |  |  | <0.001 |
| 1 | 87,062 (20%) | 75,449 (21%) | 9,587 (18%) | 2,026 (14%) |  |
| 2 | 85,806 (20%) | 73,836 (20%) | 9,757 (19%) | 2,213 (16%) |  |
| 3 | 86,465 (20%) | 73,838 (20%) | 10,076 (19%) | 2,551 (18%) |  |
| 4 | 86,281 (20%) | 72,516 (20%) | 10,770 (21%) | 2,995 (21%) |  |
| 5 | 86,142 (20%) | 69,478 (19%) | 12,258 (23%) | 4,406 (31%) |  |
| **BMI (n, (%))** |  |  |  |  | <0.001 |
| Normal | 147,018 (34%) | 132,769 (36%) | 12,923 (25%) | 1,326 (9.3%) |  |
| Obese | 96,330 (22%) | 71,177 (19%) | 17,122 (33%) | 8,031 (57%) |  |
| Overweight | 185,001 (43%) | 158,406 (43%) | 21,940 (42%) | 4,655 (33%) |  |
| Underweight | 2,337 (0.5%) | 2,047 (0.6%) | 278 (0.5%) | 12 (<0.1%) |  |
| **HbA1c, mmol/mol (median (Q1, Q3))** | 35.0 (32.6, 37.4) | 34.3 (32.2, 36.3) | 40.4 (39.5, 41.9) | 51.7 (45.6, 60.5) | <0.001 |
| **SBP, mmHg (median (Q1, Q3))** | 136 (124, 149) | 135 (124, 149) | 140 (128, 153) | 140 (130, 152) | <0.001 |
| **DBP, mmHg (median (Q1, Q3))** | 82 (75, 89) | 82 (75, 89) | 83 (76, 90) | 82 (75, 88) | <0.001 |
| **Smoking status (n, (%))** |  |  |  |  | <0.001 |
| Current smoker | 44,089 (10%) | 34,597 (9.5%) | 7,946 (15%) | 1,546 (11%) |  |
| Never smoked | 238,701 (55%) | 206,228 (56%) | 26,112 (50%) | 6,361 (45%) |  |
| Unknown | 2,107 (0.5%) | 1,561 (0.4%) | 352 (0.7%) | 194 (1.4%) |  |
| Former smoker | 147,388 (34%) | 123,165 (34%) | 18,119 (34%) | 6,104 (43%) |  |
| **Alcohol intake frequency (n, (%))** |  |  |  |  | <0.001 |
| Daily drinker | 191,127 (44%) | 167,732 (46%) | 19,459 (37%) | 3,936 (28%) |  |
| Moderate drinker | 112,131 (26%) | 95,823 (26%) | 13,101 (25%) | 3,207 (23%) |  |
| Non-drinker | 32,481 (7.5%) | 24,401 (6.7%) | 5,762 (11%) | 2,318 (16%) |  |
| Unknown | 929 (0.2%) | 630 (0.2%) | 189 (0.4%) | 110 (0.8%) |  |
| Social drinker | 95,617 (22%) | 76,965 (21%) | 14,018 (27%) | 4,634 (33%) |  |
| **Diet (n, (%))** |  |  |  |  | <0.001 |
| High | 147,875 (34%) | 125,356 (34%) | 17,644 (34%) | 4,875 (34%) |  |
| Intermediate | 168,123 (39%) | 142,543 (39%) | 20,192 (38%) | 5,388 (38%) |  |
| Low | 116,287 (27%) | 97,652 (27%) | 14,693 (28%) | 3,942 (28%) |  |
| **Sleep duration (n, (%))** |  |  |  |  | <0.001 |
| <7 h per day | 111,429 (26%) | 91,604 (25%) | 15,145 (29%) | 4,680 (33%) |  |
| >=9 h per day | 27,071 (6.3%) | 21,935 (6.0%) | 3,749 (7.1%) | 1,387 (9.8%) |  |
| 7-8 h per day | 293,340 (68%) | 251,728 (69%) | 33,548 (64%) | 8,064 (57%) |  |
| **Blood pressure-lowering medication (n, (%))** | 84,115 (19%) | 59,160 (16%) | 15,663 (30%) | 9,292 (65%) | <0.001 |
| **Oral glucose-lowering medication (n, (%))** | 10,228 (2.4%) | 0 (0%) | 0 (0%) | 9,253 (65%) | <0.001 |
| **Insulin (n, (%))** | 4,234 (1.0%) | 0 (0%) | 0 (0%) | 2,156 (15%) | <0.001 |
| **Lipid lowering medication (n, (%))** | 85,248 (20%) | 57,229 (16%) | 16,862 (32%) | 11,157 (79%) | <0.001 |
| **Family history of diabetes (n, (%))** | 89,269 (21%) | 69,310 (19%) | 13,465 (26%) | 6,494 (46%) | <0.001 |
| **Family history of dementia (n, (%))** | 50,530 (12%) | 42,458 (12%) | 6,614 (13%) | 1,458 (10%) | <0.001 |
| **PRS T2D tertile (n, (%))** |  |  |  |  | <0.001 |
| 1 | 142,617 (33%) | 128,718 (35%) | 12,410 (24%) | 1,489 (10%) |  |
| 2 | 142,617 (33%) | 122,020 (33%) | 17,203 (33%) | 3,394 (24%) |  |
| 3 | 142,616 (33%) | 111,318 (30%) | 22,425 (43%) | 8,873 (62%) |  |
| Missing | 4,435 (1.0%) | 3,495 (1.0%) | 491 (0.9%) | 449 (3.2%) |  |
| **APOE ε4 carrier, (n, (%))** |  |  |  |  | <0.001 |
| Missing | 65,676 (15%) | 54,406 (15%) | 8,750 (17%) | 2,520 (18%) |  |
| No | 259,822 (60%) | 220,118 (61%) | 31,401 (60%) | 8,303 (60%) |  |
| Yes | 103,969 (24%) | 88,862 (24%) | 12,092 (23%) | 3,015 (22%) |  |

Abbreviations: BMI, body mass index; HbA1c, glycated hemoglobin; PRS, polygenic risk score; APOE4, apolipoprotein E ε4 allele. Continuous variables are presented as median (interquartile range). Categorical variables are presented as number (percentage). Comparisons across glycaemic status groups were conducted using the Kruskal-Wallis test for continuous variables and the chi-squared test for categorical variables.

### 3.2 Primary outcomes: dementia (Fine-Gray competing risks)

As shown in *Figure 2 (a-c)*, pre-diabetes was consistently associated with higher dementia risk versus normoglycemia. For all-cause dementia, the unadjusted SHR was 1.60 (95% CI: 1.51-1.70) and remained significant after sequential adjustment (fully adjusted SHR = 1.32; 95% CI: 1.22-1.43). By dementia subtype, pre-diabetes was associated with AD (SHR = 1.30; 95% CI: 1.16-1.45) and VaD (SHR = 1.50; 95% CI: 1.28-1.76). Risks were greater for T2D, but the stepwise gradient indicates that excess dementia risk emerges already at the pre-diabetic stage.

### 3.3 Primary outcomes: stroke (Fine-Gray competing risks)

Results were similar for stroke (*Figure 2, d-f*). For all-cause stroke, pre-diabetes showed an unadjusted SHR of 1.46 (95% CI: 1.38-1.55), attenuating but persisting after full adjustment (SHR = 1.24; 95% CI: 1.16-1.33). By subtype, ischemic stroke risk was elevated (SHR = 1.24; 95% CI: 1.15-1.34), and intracerebral hemorrhage was also higher (SHR = 1.38; 95% CI: 1.18-1.62). As with dementia, risks were greater among T2D, yet the consistent excess risk in pre-diabetes indicates cerebrovascular vulnerability precedes overt diabetes.

### 3.4 Secondary outcomes: brain MRI measures (linear regression)

In fully adjusted models (*Figure 3*), pre-diabetes was associated with lower total brain volume (β =-0.28; 95% CI:-0.31 to-0.24) and higher log-WMH volume (β = 0.23; 95% CI: 0.20-0.27). T2D was associated with lower total brain volume (β =-0.31; 95% CI:-0.39 to - 0.23) and higher WMH (β = 0.17; 95% CI: 0.08-0.25). Associations were generally stronger for grey matter volume (β =-0.30 for pre-diabetes;-0.38 for T2D) and included smaller hippocampal volume in both groups. Findings were statistically significant across most MRI outcomes after correction for the false discovery rate.

**Figure 3.**
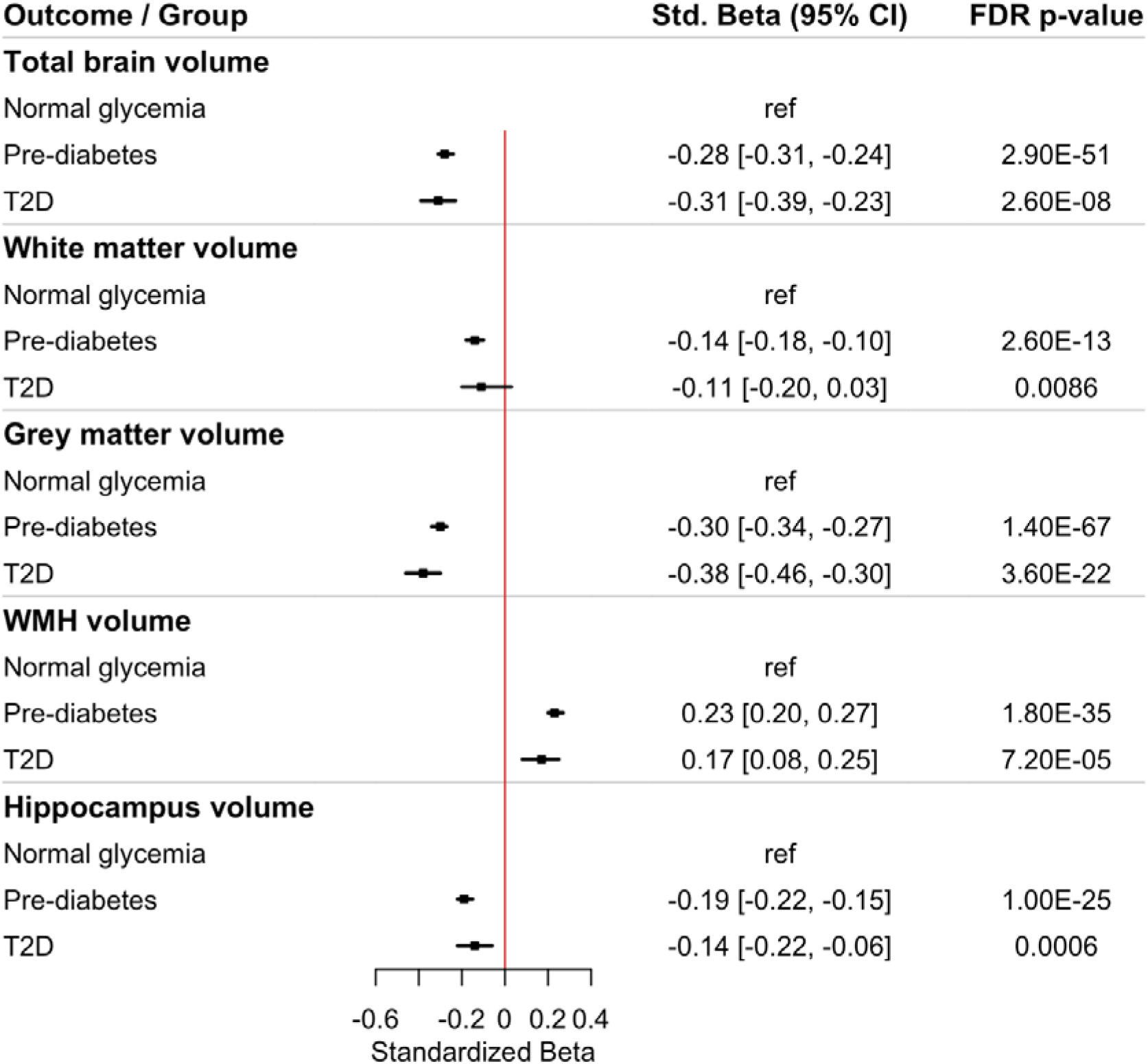
Associations between glycemia status and brain MRI measures: standardized beta estimates from fully adjusted linear models Abbreviations: T2D, type 2 diabetes; CI, confidence interval; FDR, false discovery rate; WMH, white matter hyperintensity. Model adjustments: Model 1 is crude. Model 2 is adjusted for age, gender, ethnicity, education, deprivation, diet, smoking, alcohol, and BMI. Model 3 further includes family history of diabetes, family history of dementia, and medication use of blood pressure-lowering medications, lipid-lowering medications, oral glucose-lowering medications, and insulin. Model 4 adds adjustments for T2D polygenic risk score and APOE4 carrier status. The WMH volumes are logged transferred.

### 3.5 HbA1c dose-response across outcomes (restricted cubic splines)

Restricted cubic spline analyses (*Figures 4a-4c*) revealed non-linear relationships between continuous HbA1c and outcomes: Dementia risk was relatively flat at lower HbA1c but increased above ∼40-45 mmol/mol, with a steeper rise for VaD; AD showed a modest upward trend at higher HbA1c. For stroke, including all-cause and ischemic stroke risks, rose progressively beyond ∼40-45 mmol/mol. Intracerebral hemorrhage displayed a U-shaped association, with the lowest risk at mid-range HbA1c. For brain structure, higher HbA1c was associated with lower total, grey, and white matter volumes, as well as hippocampal volumes, and higher WMH, with declines most pronounced above ∼40-45 mmol/mol.

**Figure 4.**
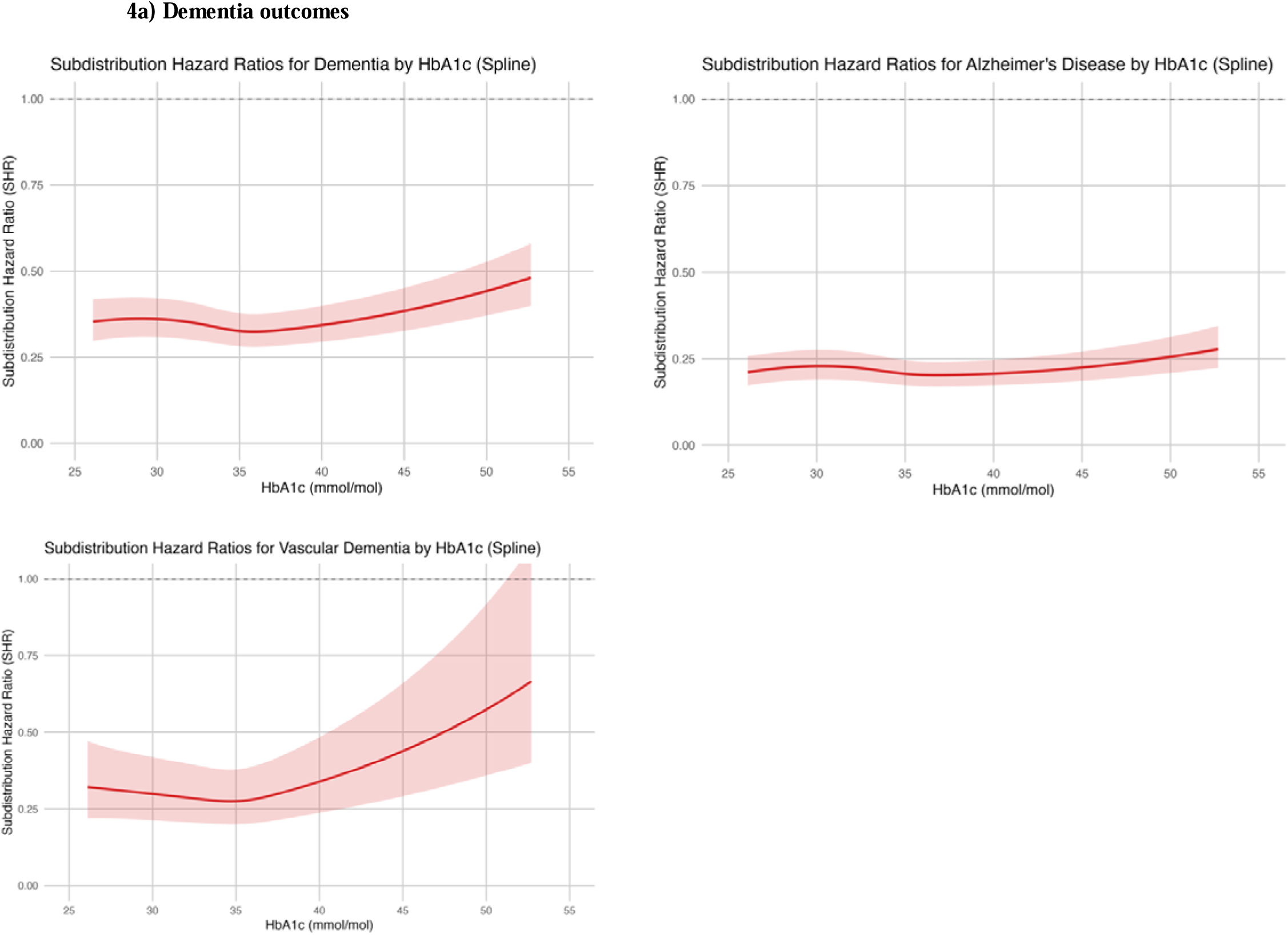
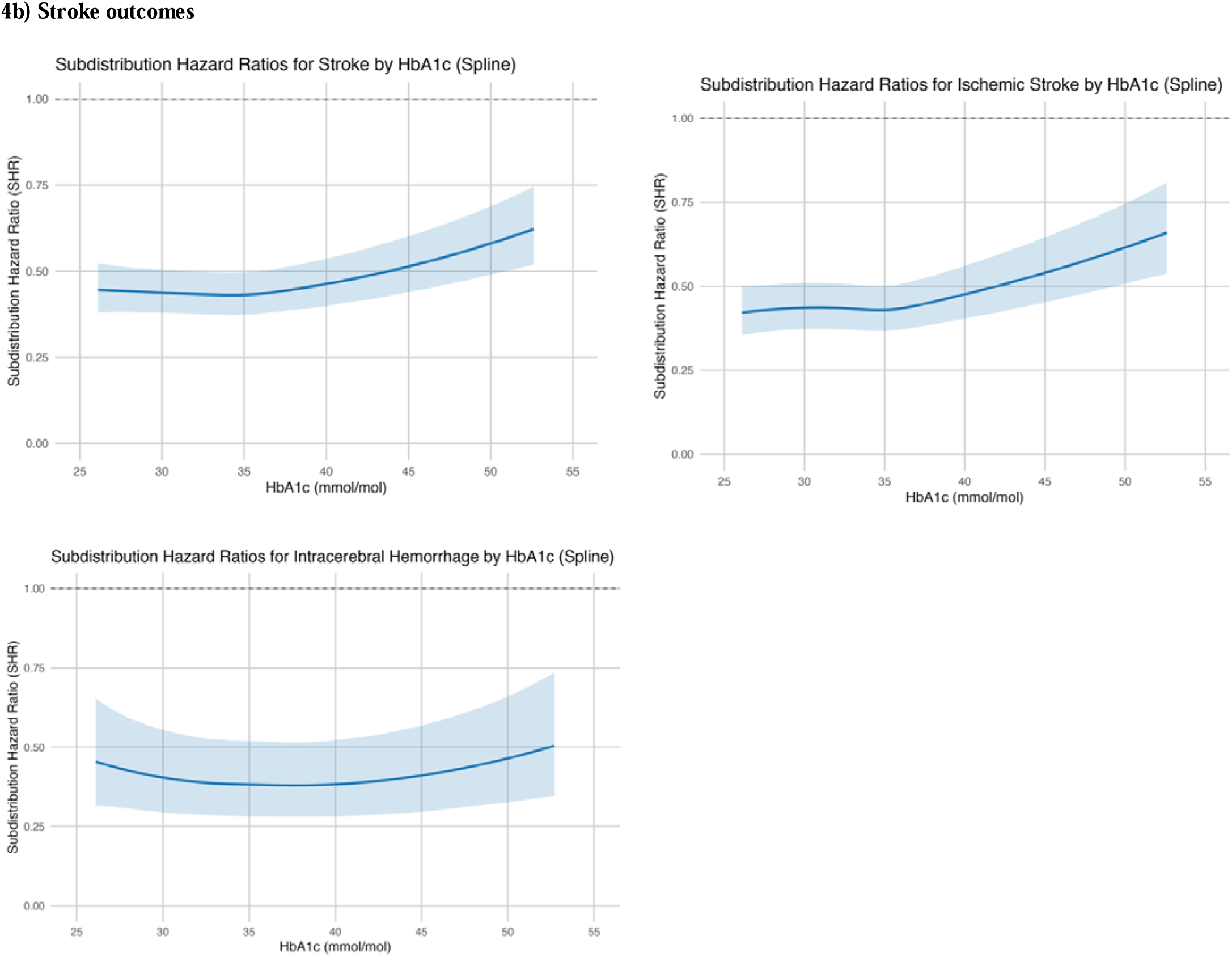
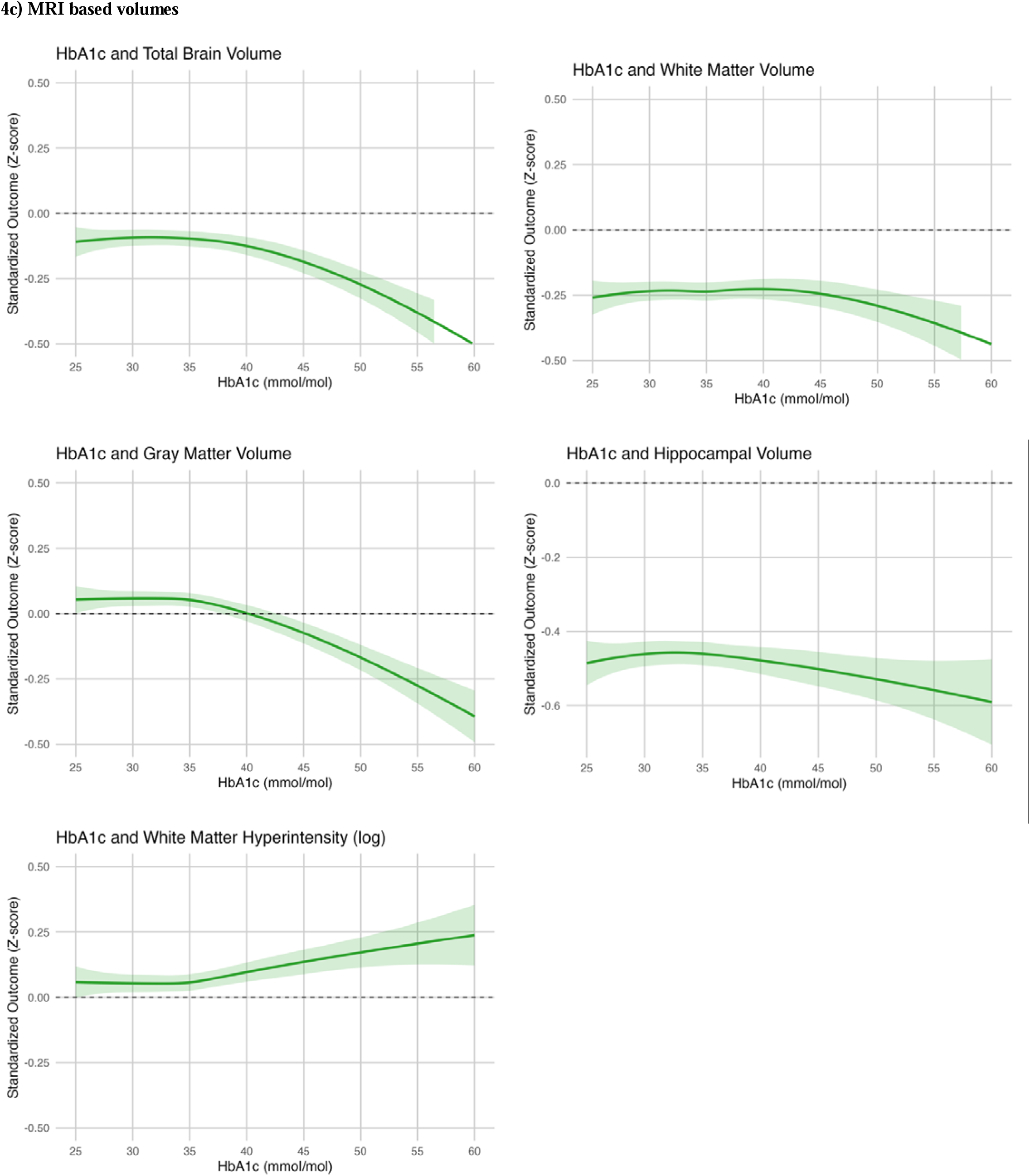
Non-linear associations between HbA1c and risk of dementia, stroke and brain MRI outcomes modeled using restricted cubic splines (Model 4). 4a) Dementia outcomes Abbreviations: HbA1c, hemoglobin A1c. HbA1c and Risk of Stroke: Stroke risk is relatively flat below ∼40 mmol/mol but increases progressively above 45 mmol/mol, with higher HbA1c associated with greater risk. HbA1c and Risk of Ischemic Stroke: The association mirrors that of overall stroke, with a clear upward trend above ∼40-45 mmol/mol, indicating that elevated HbA1c primarily drives ischemic events. HbA1c and Risk of Intracerebral Hemorrhage: The curve remains broadly flat across the HbA1c distribution, with no strong evidence of association, although confidence intervals widen at extreme values. 4c) MRI based volumes Abbreviations: HbA1c, hemoglobin A1c. Y-axes show model-predicted mean values of standardized outcomes (in SD units), adjusted for covariates. Thus, values reflect the expected deviation from the population mean outcome at a given HbA1c level, while holding other variables constant. Total brain volume: Higher HbA1c was associated with progressively lower total brain volume, with a decline of ∼0.5 SD at 60 mmol/mol compared with average levels. White matter volume: White matter volume showed a broadly similar inverse pattern, with more pronounced reductions above ∼45 mmol/mol. Gray matter volume: Gray matter volume declined steadily with increasing HbA1c, suggesting dose-dependent atrophy. Hippocampal volume: Associations with hippocampal volume were weaker; only modest reductions were seen at higher HbA1c values. White matter hyperintensity (log-transformed): In contrast, higher HbA1c was associated with greater WMH burden, with values ∼0.25 SD above the mean at 60 mmol/mol.

### 3.6 Mendelian randomization

Using robust genetic instruments (*Supplementary Tables S2-S8*), genetically predicted T2D was causally associated with higher risk of all ischemic stroke (Odds ratio (OR) = 1.14; 95% CI: 1.11-1.17; p = 5.05 × 10^-27^) and specifically with lacunar stroke (OR = 1.15; 95% CI: 1.09-1.22; p = 1.67 × 10^-7^), independent of BMI in multivariable MR. Among glycemic traits, genetically predicted HbA1c was associated with increased AD risk (OR = 1.35; 95% CI: 1.08-1.68; p = 0.008), surviving multiple-testing correction and remaining significant after BMI adjustment. For brain structure, genetically predicted T2D was inversely associated with grey matter volume (β =-0.032 SD; p = 0.005) independent of BMI. Genetically predicted HbA1c was inversely associated with hippocampal volume (β =-0.49 SD per 1 mmol/mol increase in HbA1c; p = 0.001); a suggestive association with white matter volume did not remain significant after correction. There was no consistent evidence of pleiotropy or instrument invalidity, supporting robustness. The summarized result can be found in *Table 2*.

**Table 2.** Mendelian Randomization estimates for the association between type 2 diabetes and glycemic traits with dementia and stroke subtypes, and brain MRI markers using the inverse-variance weighted method.

| Exposure | Outcome | nSNP | OR | 95% CI | p-value (adjusted) |
| --- | --- | --- | --- | --- | --- |
| T2D | Dementia | 394 | 1.04 | (0.99, 1.09) | 0.124 |
|  | AD | 356 | 0.94 | (0.83, 1.08) | 0.383 |
|  | VaD | 395 | 0.97 | (0.84, 1.13) | 0.688 |
|  | Ischemic stroke | 393 | 1.14 | (1.11, 1.17) | <0.001 |
|  | Lacunar stroke | 358 | 1.15 | (1.09, 1.22) | <0.001 |
| HbA1c | Dementia | 169 | 1.33 | (1.02, 1.74) | 0.036 |
|  | AD | 138 | 1.35 | (1.08, 1.68) | 0.008 |
|  | VaD | 169 | 0.61 | (0.32, 1.17) | 0.135 |
|  | Ischemic stroke | 158 | 1.02 | (0.86, 1.22) | 0.808 |
|  | Lacunar stroke | 143 | 1.15 | (0.80, 1.65) | 0.437 |
|  |  |  | **Beta** |  |  |
| T2D | Total brain volumes | 405 | -0.01 | (-0.03, 0.02) | 0.470 |
|  | White matter volumes | 405 | 0.02 | (-0.01, 0.04) | 0.206 |
|  | Grey matter volumes | 405 | -0.03 | (-0.05, -0.01) | 0.005 |
|  | Hippocampus volumes | 405 | 0.04 | (-0.02, 0.10) | 0.183 |
|  | WMH volumes | 405 | 0.002 | (-0.02, 0.03) | 0.834 |
| HbA1c | Total brain volumes | 174 | -0.12 | (-0.24, 0.00) | 0.056 |
|  | White matter volumes | 174 | -0.16 | (-0.29, -0.03) | 0.016 |
|  | Grey matter volumes | 174 | -0.03 | (-0.15, 0.09) | 0.595 |
|  | Hippocampus volumes | 174 | -0.49 | (-0.79, -0.19) | 0.001 |
|  | WMH volumes | 174 | 0.008 | (-0.11, 0.13) | 0.901 |

Abbreviations: T2D, type 2 diabetes; OR, odd ratio; HbA1c, hemoglobin A1c; AD, Alzheimer's disease; VaD, vascular dementia; WMH, white matter hyperintensities; CI, confidence interval; SNP, single nucleotide polymorphism
Odds ratios are shown for genetically predicted T2D, HbA1 with dementia, AD, and VaD.

## Discussion

In this large, population-based study integrating prospective observational analyses with MR, pre-diabetic dysglycemia was consistently associated with higher risks of dementia, stroke, and adverse brain structural profiles. Across the HbA1c continuum, risk elevations were already evident within the pre-diabetic range and increased further with progression to T2D. These convergent findings indicate that an increased risk of dementia and stroke can emerge at sub-diabetic glycemic levels, highlighting pre-diabetes as a critical stage for risk stratification and prevention.

For dementia, pre-diabetes was independently associated with elevated risks of all-cause dementia and its subtypes after extensive adjustment. Effect sizes were modest to moderate but consistent: fully adjusted SHR were approximately 1.32 for all-cause dementia, 1.30 for AD, and 1.50 for VaD. Although risks were higher among individuals with T2D, the stepwise gradient observed across glycemic categories indicates that excess dementia risk is present before overt diabetes. Restricted cubic spline analyses further showed that dementia risk rose from around 40 mmol/mol HbA1c, below the diagnostic threshold for diabetes, with the steepest increases for VaD, consistent with a vascular contribution to cognitive decline rather than a discrete threshold effect.

Stroke analyses reinforced the vascular implications of early dysglycemia. Pre-diabetes was associated with higher risks of all-cause stroke, ischemic stroke (fully adjusted SHR ∼1.24), and intracerebral hemorrhage (ICH; ∼1.38). Spline models indicated monotonic increases for ischemic stroke beyond about 40 mmol/mol, while ICH displayed a U-shaped relationship with the lowest risk at mid-range HbA1c. These subtype-specific patterns suggest distinct pathophysiological pathways and underscore that cerebrovascular vulnerability is detectable already in pre-diabetes.

Neuroimaging results provided structural correlates of these clinical associations. Compared with normoglycemia, pre-diabetes was associated with smaller total and grey matter volumes, smaller hippocampal volume, and greater WMH burden (22, 23); effects were generally larger in T2D but were not confined to overt diabetes. Findings for white matter volume were less consistent and did not uniformly survive multiple-comparison correction, whereas WMH showed clear and robust increases. Continuous HbA1c modeling indicated that structural changes became more pronounced from approximately 40 mmol/mol, reinforcing the clinical relevance of moderate glycemic elevations.

MR analyses offered complementary causal support that mapped onto the observational patterns. Genetically predicted T2D was associated with higher risks of ischemic and lacunar stroke, independent of BMI, while genetically predicted HbA1c was associated with higher risk of AD and with smaller hippocampal volume. These results reduce concerns about residual confounding and reverse causality for these specific pathways and highlight both macro-and microvascular as well as neurodegenerative mechanisms linking dysglycemia to adverse brain outcomes.

Several biological mechanisms may underlie these associations. Chronic hyperglycemia and insulin resistance can promote endothelial dysfunction, impaired cerebral autoregulation, and microangiopathy; together, these processes may increase susceptibility to both ischemic and hemorrhagic events (24, 25). Glucotoxicity may also drive oxidative stress and low-grade inflammation, accelerating the formation of advanced glycation end-products (AGEs) (26), mechanisms implicated in neurodegeneration and β-amyloid pathology (27–29). Increased WMH burden and smaller grey/hippocampal volumes align with small-vessel disease and neurodegenerative vulnerability (30–33), providing a plausible substrate for the observed clinical risks.

This study has important strengths. The large, well-characterized cohort with long follow-up enabled precise estimation of associations across dementia subtypes, stroke subtypes, and neuroimaging markers. Modeling glycemia both categorically and continuously (via splines) provided a nuanced dose-response view, and MR analyses offered complementary causal inference, including multivariable MR adjusting for BMI and sensitivity analyses to probe pleiotropy.

Limitations, however, merit consideration. Despite extensive adjustment, residual confounding cannot be excluded, and routine-record ascertainment may introduce outcome misclassification. UK Biobank’s healthy-volunteer bias and predominantly European ancestry limit generalizability. HbA1c was measured once at baseline; regression-dilution bias and unmeasured glycemic variability could attenuate estimates. Progression from pre-diabetes to T2D was not modeled as a time-updated exposure, which may underestimate pre-diabetes–related risk accrued before conversion. Fully adjusted models include cardiometabolic factors and medications that may act as mediators; over-adjustment could bias associations toward the null. Imaging analyses were restricted to a subset and may be susceptible to selection biases, although the results were robust after false discovery rate correction. Another limitation is the absence of GWAS specifically focused on pre-diabetes, which precluded the construction of valid instruments for this glycemic category in the MR analyses. Future GWAS dedicated to intermediate glycemic states would enable more comprehensive causal inference across the glycemic spectrum.

Taken together, these findings provide consistent evidence that even modest glycemic elevations, below current diagnostic thresholds, are linked to increased risks of dementia and stroke and to structural brain changes. By combining prospective epidemiology, flexible dose–response modeling, and genetic causal inference, the study strengthens the plausibility of early neurovascular and neurodegenerative effects of dysglycemia and positions the brain as a target organ in metabolic health.

### Clinical implications and future studies

Clinically, pre-diabetes should be recognized as a state of heightened neurocognitive as well as cardiometabolic risk. Routine HbA1c assessment can help in stratifying risk for dementia and stroke, and earlier intervention, through lifestyle modification and, where appropriate, pharmacotherapy, may help preserve cognitive function and brain structure. Incorporating brain health considerations into metabolic care, especially for older adults and those with longstanding dysglycemia, is warranted. Future research should test whether improving glycemic control in pre-diabetes attenuates or reverses imaging markers and reduces clinical events, ideally via randomized trials. Longitudinal imaging studies can clarify temporal dynamics and mechanisms, and larger MR analyses stratified by age, sex, and ancestry can refine causal pathways and identify vulnerable subgroups. Prioritizing brain health within diabetes prevention and management, beginning in pre-diabetes, may improve long-term outcomes across cognitive and cardiovascular domains.

## Data Availability

All data produced in the present study are available upon reasonable request to the authors.

## Author Contributions

S.H., F.A. and G.J.B. conceived the idea presented. S.H. conducted all statistical analyses under the supervision of F.A. and G.J.B.. S.H. drafted the manuscript, with F.A. providing critical revisions and feedback. All authors have reviewed and approved the final version of the manuscript.

## Availability of data and materials

This research was conducted using the UK Biobank resource under application number 100993. The UK Biobank data are available to bona fide researchers upon application and approval by UK Biobank (www.ukbiobank.ac.uk). Restrictions apply to the availability of these data, which were used under license for the current study and are not publicly available. The GWAS summary statistics used in this study are publicly available from the following sources: https://www.ebi.ac.uk/gwas/ (accessed on 1st Dec 2024). Data on HbA1c have been contributed by MAGIC investigators and have been downloaded from www.magicinvestigators.org.

